# Network analysis of England’s single parent household COVID-19 control policy impact: a proof-of-concept study

**DOI:** 10.1101/2021.10.26.21265363

**Authors:** N.L. Edelman, P. Simon, J.A. Cassell, I.Z. Kiss

## Abstract

Lockdowns have been a key infection control measure for many countries during the COVID-19 pandemic. In England’s first lockdown, children of single parent households (SPHs) were permitted to move between parental homes. By the second lockdown, SPH support bubbles between households were also permitted, enabling larger within-household networks. We investigated the combined impact of these approaches on household transmission dynamics, to inform policymaking for control and support mechanisms in a respiratory pandemic context.

This network modelling study applied percolation theory to a base model of SPHs constructed with population survey estimates of SPH family size. To explore putative impact, varying estimates were applied regarding extent of bubbling and proportion of Different-parentage SPHs (DSPHs) (in which children do not share both the same parents). Results indicate that the formation of giant components (in which Covid-19 household transmission accelerates) are more contingent on DSPHs than on formation of bubbles between SPHs; and that bubbling with another SPH will accelerate giant component formation where one or both are DSPHs. Public health guidance should include supportive measures that mitigate the increased transmission risk afforded by support bubbling among DSPHs. Future network, mathematical and epidemiological studies should examine both independent and combined impact of policies.

## Introduction

The COVID-19 pandemic has had a devastating impact around the World; with politicians implementing population infectious disease control measures including ‘lockdowns’, defined by the Cambridge dictionary as: *‘a period of time in which people are not allowed to leave their homes or travel freely, because of a dangerous disease’*.

After reporting its first case in January 2020, the UK has gone on to have one of the highest excess death rates in Europe (7.7% above the five-year average by the end of that year) (1). The first national lockdown was imposed on 23^rd^ March 2020, and eased through June/July of that year. The ‘Stay at Home’ rules exempted children from ‘single parent households’ (SPHs) who routinely stayed with or visited both parents such that they could continue to do so. After a flattening and decline in the epidemic curve through the summer a second English lockdown was imposed on 3^rd^ November 2020 and re-introduced on 6th January 2021 after being briefly lifted through December using a tiered system of restrictions. Other social-distancing measures implemented to varying degrees across the UK and other countries include disallowing and/or limiting individuals being inside others’ homes.

Following the first lockdown evidence emerged of the negative impact of social-distancing on mental well-being (2). Social support was found to have an important protective effect against this (3). Therefore, from September 2020 some households in England were permitted to form a ‘support bubble’ with another household (4). SPHs were among these designated household types, defined as *‘a single adult living with one or more children who are under the age of 18 or were under that age on 12 June 2020’* (4). The Office for National Statistics (ONS) estimates there were 2.9 million lone parent families (LPF) in the UK in 2020 (14.7% of all families). This calculation attributes each child to one household based on which parent receives the child maintenance. In 86% of LPFs the child(ren) resides predominantly with the mother (5). Estimates of LPF size differ greatly. ONS Families and Household data for 2018 found that 55% of LPFs had one child, 32% had two and 13% comprised three or more (<18 years of age); while 2020 data found 30.17% had one child, 37.34% had two and 32.49% had three or more. Amongst LPFs there is no available data on discordant parentage (in which the children of that household do not share all the same parents/caregivers), although US data suggests that 28% of all women with two or more children had those children by more than one father (6).

The risk of household transmission of SARS-Cov-2 is estimated to be ten times greater than for transmission via non-household contacts (7); and thought to account for 70% of all transmission (8). The household secondary attack rate (the percentage of contacts of an index case who are also infected) has been estimated at 16.6% - varying by age, relationship type, presence of symptoms and number of household members (9). Network analysis has aided understanding of transmission dynamics where household living arrangements are combined with movement of actors between homes; a recent study of domiciliary care demonstrating the impact on COVID-19 transmission of that movement (10).

Infectious disease transmission amongst individuals connected via a contact network, can be mapped using percolation theory (11) (12). Put simply, the transmission of an epidemic between network nodes requires ‘activation’ of links along which infection has spread. Keeping such links and discarding all other, also referred to as ‘bond percolation’, results in a network with fewer links, since not all existing links will transmit. The severity of an epidemic is directly related to the size of connected components (i.e. a subset of nodes where any two nodes are connected in both directions) in this sparser network. A giant component is a subset of nodes (e.g. adults and children in SPHs) such that any two nodes (people) in this subset can be connected using the available links (such as those afforded from OHL and/or support bubbling); and where the number of nodes in this subset scale with the size of the full network. Where a giant component is formed, a severe epidemic is likely as the infection can ‘percolate’ through the network. Conversely, if the sparser network comprises many disconnected components, the likelihood of a severe epidemic is small. In percolation theory, one is interested in the point at which the extent of connectivity between people leads to the formation of a ‘giant component’. From a public health perspective, this is particularly valuable in identifying the critical percolation threshold at which an infection (such as SARS-CoV-2) is likely to affect a large proportion of individuals. Percolation theory has already been applied to investigate putative impact on COVID-19 transmission of forming household bubbles, finding that reduced contact outside the bubble mitigated the transmission impact of bubbles with more than two members (13). Network analysis has also explored the potential impact on COVID-19 transmission of ‘contact clustering’ in social bubbles (as part of an imagined exit strategy), similarly finding negligible impact where those bubbles remained contained (14). However, both studies assume exclusivity between bubbling households, not accounting for domiciliary movement between households such as that afforded by care workers or the children of SPHs. To the authors’ knowledge, no studies have investigated the cumulative impact on COVID-19 transmission of SPH support bubbles in the context of SPH child movement between parental residences.

This proof-of-concept study aimed to examine the impact of two mechanisms by which a network of SPHs in which children are spending time with parents who live in different households can become more connected, leading to larger giant components with a higher probability of a large outbreak. (Here we use the term “parent” to include all primary care givers regularly resident in the same household with the child). The first mechanism was the extent of discordant parentage within SPHs (i.e., SPHs that comprise two or more children who only share one parent (having different parents in other SPHs with whom they also stay with regularly). We define households of this type as Discordant-Parentage Single Parent Households (DSPHs); for example, a DSPH might comprise a foster carer, Child A and Child B (who are not related and each stay with their own different paternal grandmother regularly). The second mechanism was the extent of bubble formation between one SPH and another SPH (that does not include the other parent of any of the offspring). For example, SPH1 may bubble with SPH2 while the two offspring of SPH1 each have a parent they stay with regularly who reside in SPH3 and SPH4 respectively rather than in SPH2 with whom the bubble is formed. Investigation of this issue offers important insights for our understanding of Covid-19 household transmission rates and for future decisions about the deployment of multiple and co-occurring infection control measures concerning contact between households in response to the COVID-19 pandemic as it progresses.

## Methods

For the purposes of this study we defined a single parent as a parent (under the definition above) who does not live with the other primary caregiver of their child or children, irrespective of whether they are living with a new partner or not. We therefore defined a single parent household (SPH) as a household in which a single parent (using our definition) resides with one or more of children at least 10% of the time. Thus, a child who stays with each parent at least one night in every fortnight is considered a member of two SPHs. This can be seen in the top-left panel of Figure 1 where for example a child (blue dot) is connected to two parents (green dots), where the parents themselves are not connected. As the study aimed to produce a static model we henceforth use the term ‘offspring household linkage’ to denote the movement of children between the two SPHs in which their parents’ reside. We hypothesized that the combined effects of SPHs bubbling with each other *in addition to* offspring household linkage of SPHs outside of those bubbles (i.e. children alternating time with each parent) would contribute to Covid-19 household transmission.

**Figure 1:**
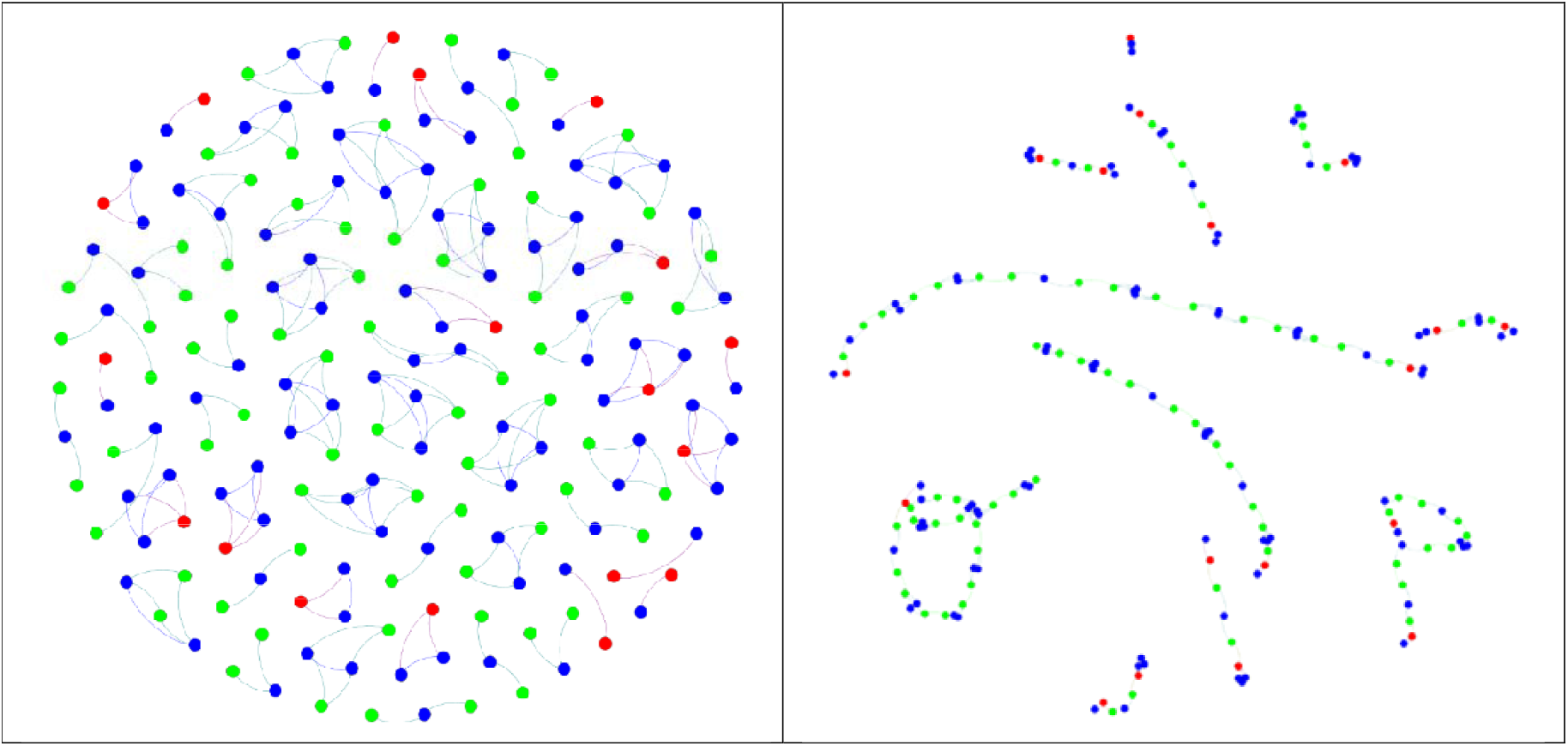
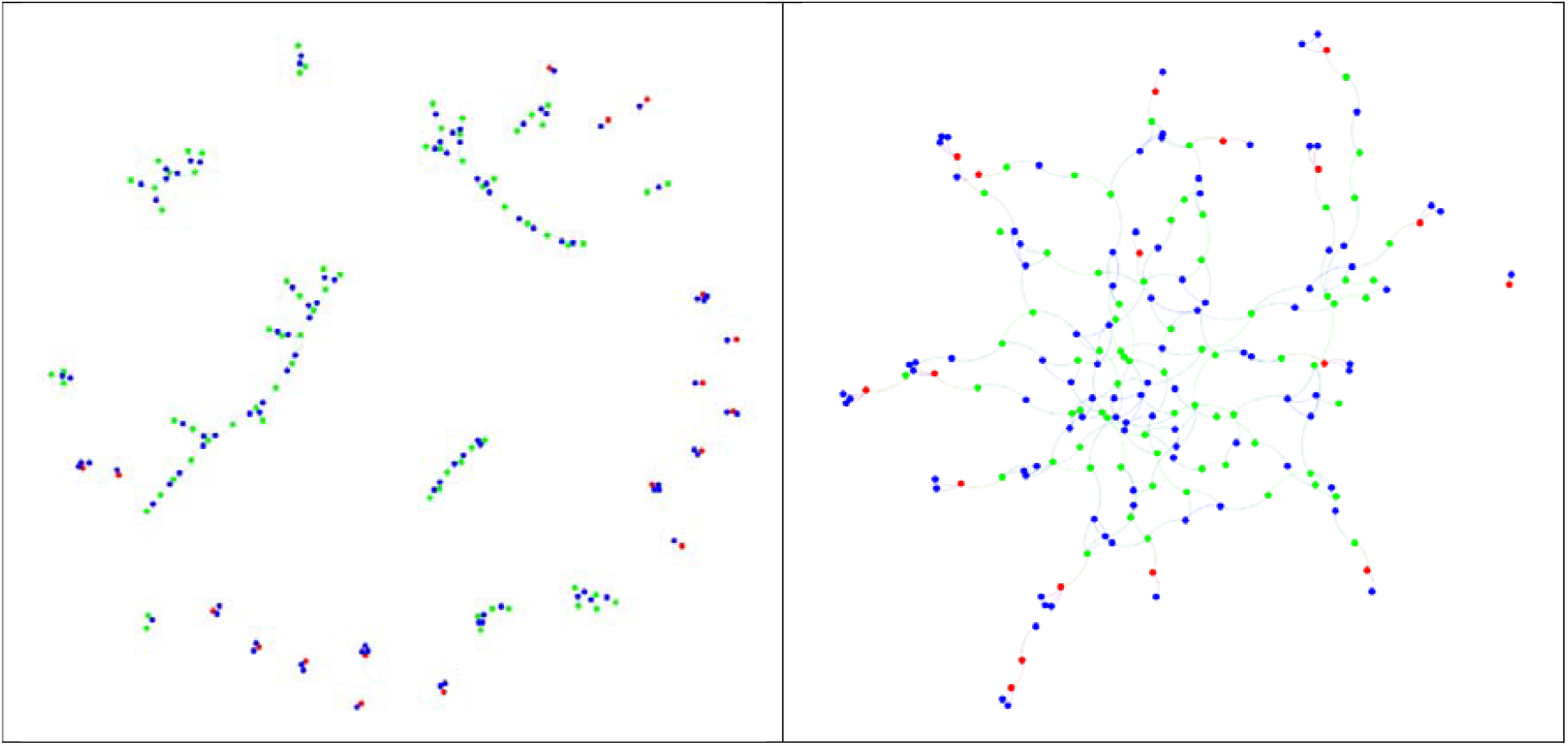
Plots of ‘toy’ contact networks (representing a reduced number of nodes for visual illustration of N=186) with different levels of DSPHs and Bubbling. (**Top-left**) Baseline model without any discordant-parentage and without any bubbling, (**Top-right**) bubbling only. (**Bottom-left**) discordant-parentage only, and (Bottom-right) both discordant-parentage and bubbling. Nodes are colour code as follow: Blue dots – children, Green dots – parents in households where both parents retain contact with offspring, and Red dots – parents who are the sole parent having regular contact with their offspring.

We further hypothesized that where a SPH is a DSPH (comprising two or more children who have parents in other SPHs with whom they also stay with regularly as described above), this would additionally contribute to network transmission and the formation of a giant component. We defined households of this type as Discordant-Parentage Single Parent Households (DSPHs). These definitions of SPH, DSPH and Offspring Household Linkage (OHL) support the analysis of COVID-19 transmission by placing focus on children spending time with parents in different SPHs and on eligibility for support bubble formation, rather than on legal residency of the child or current relationship status of the parent. In this study we developed a simple mechanistic network model upon which to explore the impact of DSPHs and bubbling as mechanisms that that can increase the connectivity in the network and thus the risk of larger outbreaks.

Relevant parameters for base model construction were drawn from population survey estimates, where these existed. UK data on rates of parent contact among dependent children of separated or divorced parents is difficult to obtain. A 2013 survey of non-resident fathers found that 13% reported no contact with their child with 59% reporting contact at least once a week; this data did not specify whether that contact was face-to-face but 49% reported their children staying with them on a weekly basis (15). An earlier 2007 study using ONS data found that approximately 35% children in SPHs stayed with both parents on a weekly basis with a further 25% staying less than once a week but more than once a month (16). We used this data as context, developing our model with the assumption that 80% of children would spend time at both their parents’ SPHs once a fortnight or more frequently.

Given the variation in distribution of number of children in SPHs (as reported in the Introduction) and the lack of direct comparability between the ONS definition of lone parent families and our own definition of SPHs we used parameter estimates representing mid-points such that 42% of SPHs had one child, 35% had two children and 23% had three or more.

We therefore defined a base model such that the number of SPHs with one, two and three children is 420, 350 and 230, respectively. To account for circumstances where contact between one parent and their offspring has ceased, the model was constructed such that 20% of all SPHs did not have an OHL to another SPH. Such households are visible in the top-left panel of Figure 1 where parents are denoted by red dots and all children are represented by blue dots. These are in line with the figures reported above. These offspring linkages (i.e. the connections between SPHs that arise from children staying in each parent’s SPH at least once a fortnight) are hard-wired into the model on ethical grounds. I.e., this was considered a ‘non-negotiable’ aspect of public health directives because it would be unethical to prevent children from visiting or staying with both parents regardless of the modelled impact on COVID-19 transmission.

The network is constructed by first focusing on the 80% of the SPHs. Knowing these numbers, say *N*_*SPH*1_, *N*_*SPH*2_ and *N*_*SPH*3_, determine the number of children in the network at this stage, that is *N*_*CH*_ = (*N*_*SPH*1_, + 2 × *N*_*SPH*2_, + 3 × *N*_*SPH*3_)/2. A proportion of *r*_l_, *r*_2_ and 1 − (*r*_l_ + *r*_2_)of the children are then allocated to SPHs with one, two and three children, respectively. **This is done proportionally to the number of stubs starting from SPHs, that is** 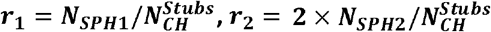 **and** 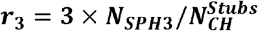, **where** 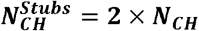. **This means that for example, *r***_**1**_ **× *N***_***CH***_ **children will be allocated to SPHs with one child, and the same approach is applied to SPHs with two and three children. At this point, all children will have exactly one spare stub remaining, that is the stub needing a second parent. Equally, there will be parents with no children allocated to them at this point**. This leaves us with a good degree of flexibility to vary the discordant-parentage in SPHs. E.g., one can impose that a stub from a child already allocated to a SPH connects to a parent with one single stub, i.e. to a parent who is in the pool of SPHs with one child. Conversely, the spare stub from this same child could be connected to a spare stub from a parent in the pool of SPHs with two or three children. The degree to which DSPHs is enforced is captured by the probability *p*_*mp*_, where a value of zero means that all spare stubs from children in SPHs with one, two or three children are allocated to parents that are in the pool of SPHs with one, two or three children, respectively. When this probability is close to one, the degree of MPSPHs is pushed to maximum within the constraints of being able to construct the network.

This process is followed by supplementing the network with an extra 20% of SPHs (for all three types of SPHs) with one parent only. These are isolated fully connected ‘cliques’ comprising two, three or four nodes, respectively (see Fig. 1 top-left panel) with parents (red dots) and children (blue dots). Such households do not contribute to the extent of DSPHs.

Next, we form bubbles between pairs of parents that do not share offspring. All permissible pairs are chosen at random and connected with probability *p*_*b*_, i.e. the probability of bubble formation across a given pair.

Once the network model is implemented, we vary (*p*_*mp*_, *p*_*b*_) and measure the size of the giant component across many realisations. We also measure the average degree in the network and consider the distribution of connected components to understand how the giant component emerges.

## Results

First, we demonstrate the effect of the two mechanisms by explicitly plotting the contact network for networks of relatively small size, in this case individuals.

Figure 1 top left-hand panel depicts the baseline model. The network consists of isolated clusters where children (blue dots) are connected to siblings and to one (red nodes) or two (green nodes) parents. Figure 1 bottom-left panel illustrates connectivity from DSPHs alone, and in the top-right panel bubbling alone. Greatest connectivity occurs where DSPHs and bubbling are combined (bottom-right panel). Bubbling alone forms multiple chains of smaller sizes (top-right panel) from which emergence of a large chain is unlikely. However, when discordant-parentage is more common, the opportunities for linkage creation between SPHs increase. For example, three children in a SPH with their father may have three different mothers, each of whom is a part of another SPH, creating more chances to connect up isolated chains of SPHs. These trends are clearer in the simulations performed on larger networks (Figure 2).

**Figure 2:**
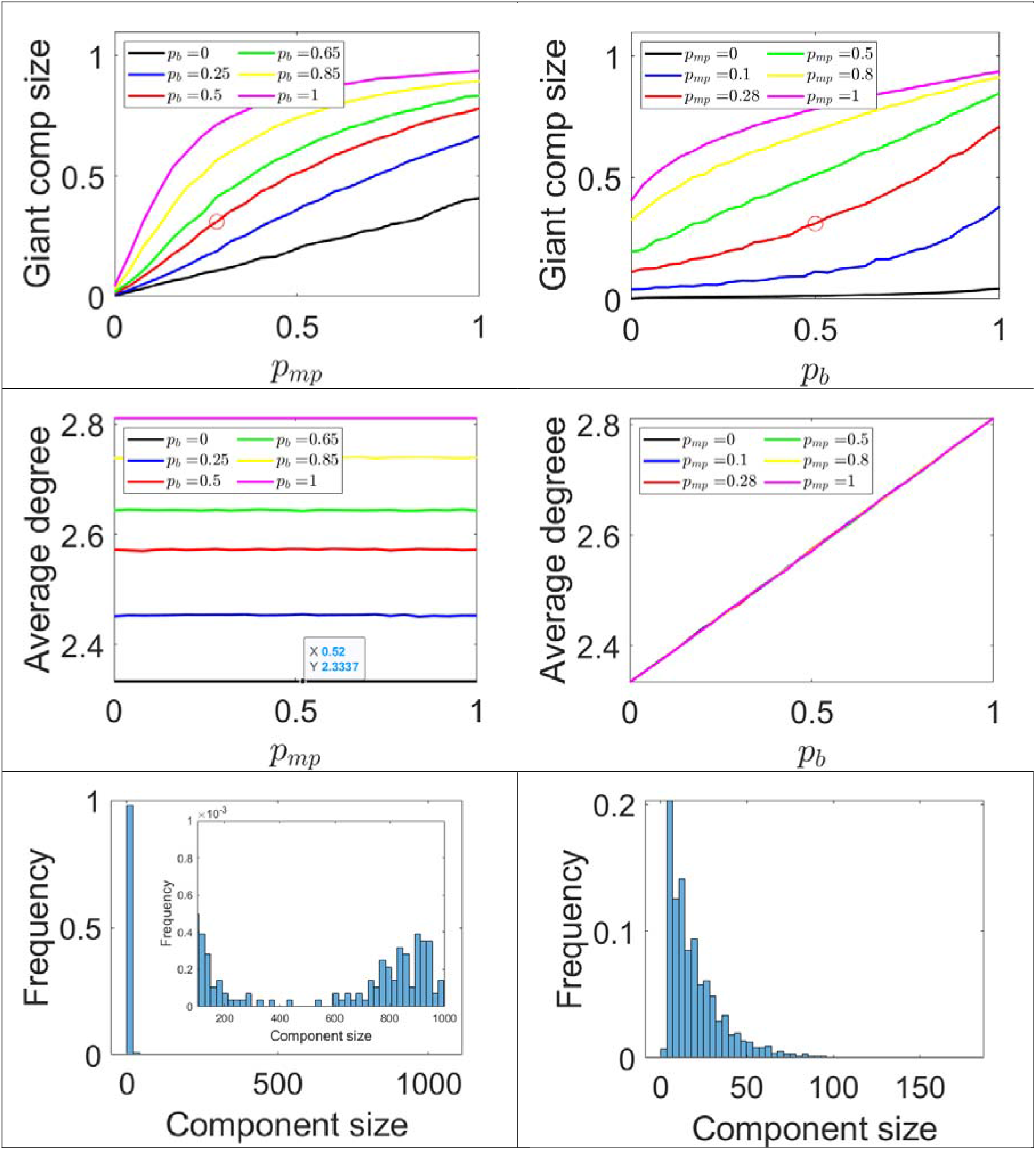
Plots of the giant component size (top row), average degree (middle row) and distribution of connected component sizes (bottom row) as a function of increasing levels of DSPHs (left column) and bubbling (right column). The last row gives the distribution of component size for (*p*_*mp*_, *p*_*b*_) = (1,0) (left panel) and (*p*_*mp*_, *p*_*b*_) = 0,1 (right panel). The red circles in the top row correspond to (*p*_*mp*_, *p*_*b*_) = (0.28,0.5), values that have been observed in practice.

We now move on to report on results run on networks of N=2086 nodes (this seemingly arbitrary number arises as SPHs with one, two and three children and their numbers lead to some conditions that are necessary to construct the network). We studied the full spectrum of values, with special focus on which seems to be close to some of the estimates in the literature described above. These demonstrate how several networks were created with a given parameter set and the measured values averaged across these realizations.

The plots in Figure 2 demonstrate the incremental impact of the two mechanisms by which the contact network can lead to the formation of a larger giant component and thus a higher probability of a large outbreak. Plots in the left-hand column depict the probability of SPHs of two or more children comprising Discordant-Parentage SPHs, described by the value, and assessing the impact of this on network connection for various fixed probabilities of bubble formation () (depicted with different coloured lines). Plots in the right-hand column depict the probability of SPHs forming bubbles with other SPHs (that do not include the other parent of that SPH’s offspring), described by the value (*p*_*b*_) and assessing the impact of this on network connections for various fixed probabilities of DSPHs (*p*_*mp*_) (depicted again with coloured lines).

The top row of Figure 2 illustrates that the growth of the giant component is much slower for bubbling without DSPHs than for DSPHs without bubbling. DSPHs create bigger components such that addition of bubbling events allows for a giant component to emerge. This is clearly visible if we inspect the black curves in top row of Figure 2. It appears that bubbling by non-discordant parentage SPHs forms a giant component only very slowly, with many smaller components emerging that nonetheless do not easily percolate into a single giant component. However, as *p*_*mp*_ increase, and with no bubbling, the giant component grows faster. This is despite the average number of links staying constant under various values of *p*_*mp*_ and increasing as *p*_*b*_ increases, see middle row of Figure 2. The intuition behind this is provide by looking at the bottom row of Figure 2. It is clear that as *p*_*mp*_ approaches its maximum value a giant component can emerge. This is not the case for value of *p*_*b*_ close to 1, where all components remain of small size.

Figure 2 includes a point estimate (see red circles) that represents the most likely UK scenario regarding prevalence of DSPHs and of bubbling. This point estimate assumes that 28% of SPHs comprise two children or more are DSPHs (based on US data and making the conservative assumption in the case of three or more children each child would not have a different other parent), and that 50% of SPHs would form a support bubble. The growth of the giant component is faster as the level of MPSHPs increases compared to when bubbling increases. Furthermore, the lines in the top-left panel correspond to networks with the same average degree. On the one hand, this is because the level of MPSPHs does not change the number of links in the network; it simply re-distributes links. However, when bubbling is acting then the network gains more links. On the other hand, this means that the same number of links can be distributed in order to increase or decrease the size of the giant component. In the top-right panel, it is evident that fixing *p*_*b*_ *=* 0.5 leads to an average degree of about 2.55 but with giant component sizes ranging from 0.1 to 0.8 approximatively.

## Discussion

This proof-of-concept study demonstrates how support bubbles between SPHs has little impact on formation of giant components that may cause Covid-19 outbreaks, except where one or more are DSPHs as offspring household linkages from DSPHs have a greater impact on giant component formation than does support bubbling with another SPH. The cumulative effect of DSPHs forming bubbles with other SPHs or DSPHs likely speeds up the formation of giant components through which COVID-19 transmission would occur.

This is the first study to model the combined effects of two SPH-related UK Covid-19 infection control measures; examining the added impact of bubble formation *between* SPHs against a backdrop of SPH network linkage created by OHL. It used prevalence estimates of SPH number of children and rates of contact with both parents for children of SPHs. In the absence of good estimates for rates of discordant parentage and extent of bubbling between SPHs, the study design allowed exploration of the variable impact of each on giant component formation – modelling separate and combined impacts.

## Limitations

This study modelled connectivity between individuals in SPHs, rather than Covid-19 transmission itself. As a proof-of-concept study, this work did not take account of *all* variables likely to affect household transmission and worked under certain hypotheses concerning family size and constitution. For example, distribution of offspring age was not accounted for, even though younger children are less likely to confer transmission (17) while older children are more likely to lose contact with their fathers, which would reduce OHL. Nor did we account for variation in the amount or frequency of time that a child spent with each parent.

In the interests of simplicity, our models did not account for the additional effect of childcare bubbling (open to all households with children aged fourteen years or younger, but with close contact minimised). The prevalence of this is not well documented and likely to have variable impact on transmission depending on the extent of contact; Public Health England guidance advised that a SPH could meet simultaneously with its childcare bubble and support bubble). As this study focused on SPH connectivity rather than SPH-related Covid-19 transmission itself, the models did not account for school-related infection control measures - when schools are open the differential impact of DSPHs forming support bubbles may well be negligible.

Use of real-life data to test our hypothesis and validate the models is not currently possible as surveillance has not captured uptake and type of support bubbles. Nor do population surveys adequately capture the size and composition of SPHs and movement of offspring between them. Surveillance data during health emergencies needs to better capture uptake of policies to inform decision-making. This study demonstrates the importance of scientists and policy makers considering the potential impact of not only *individual* infection control measures but their potential *combined* effects. This supports not only accurate assessment of *overall* impact, but also identification of differential impacts that require mitigations to better support specific sub-populations. The findings indicate that support bubbles generally make little contribution to Covid-19 transmission. However, potential for significant contribution to transmission is greater with bubbling involving DSPHs.

Support bubbles are an important strategy for social and psychological support when social interactions is restricted; the findings suggest that additional support strategies may be needed to mitigate increased risk of transmission from support bubbling among DSPHs. Importantly, low income is associated with dense living accommodation (18), and also with both parental separation and higher numbers of children (for women, who are more often the primary carer) (19). Lower income has also been found to be associated with reluctance to test for Covid-19 with significantly lower testing rates in areas of economic deprivation, widely attributed to the economic impact of self-isolation where individuals are on zero-hour contracts or in other unstable employment (20). Strategies are therefore warranted to encourage frequent testing, including co-produced health promotion messaging and adequate financial recompense for those self-isolating. The increased potential for transmission where DSPHs bubble, also points to the need for effective mitigations against household transmission - such as ventilation - that recognise the structural vulnerability of these families.

## Bullet point summary

- Support bubbles formed between single parent households are unlikely to contribute significantly to household transmission except where these households comprise children of different parentage who are each spending time with both of their parents
- SPHs that comprise children of different parentage who are moving between parental homes, contribute more to Covid-19 transmission acceleration than support bubble formation with another single parent household
- Measures to support frequent testing are warranted in single parent households comprising children of different parentage, such as co-produced health promotion and more substantial financial recompense to enable self-isolation following a positive result
- Infectious disease control strategies should be assessed regarding both individual and combined effects in order to identify where additional supportive measures are needed to mitigate interacting effects

## Data Availability

All data produced in the present study are available upon reasonable request to the authors

## Author contributions

NE conceived the initial idea, consulted on the analysis design, and drafted the paper.

PS contributed to analysis design and execution and to publication drafts.

JC advised on study design and contributed to publication drafts.

IK led the design, implemented code, analysed results and wrote parts of the original draft.

Data availability

The data that support the findings are available by direct communication with Professor Istvan Kiss in line with University of Sussex policy.

